# Topological data analysis identifies distinct biomarker phenotypes during the ‘inflammatory’ phase of COVID-19

**DOI:** 10.1101/2021.12.25.21268206

**Authors:** Paul W. Blair, Joost Brandsma, Josh Chenoweth, Stephanie A. Richard, Nusrat J. Epsi, Rittal Mehta, Deborah Striegel, Emily G. Clemens, David A. Lindholm, Ryan C. Maves, Derek T. Larson, Katrin Mende, Rhonda E. Colombo, Anuradha Ganesan, Tahaniyat Lalani, Christopher J Colombo, Allison A. Malloy, Andrew L. Snow, Kevin L. Schully, Charlotte Lanteri, Mark P. Simons, John S. Dumler, David Tribble, Timothy Burgess, Simon Pollett, Brian K. Agan, Danielle V. Clark, the EPICC COVID-19 Cohort Study Group

**Affiliations:** The Henry M. Jackson Foundation for the Advancement of Military Medicine, Inc., Bethesda, MD; Department of Pathology, Uniformed Services University, Bethesda, MD; Infectious Disease Clinical Research Program, Department of Preventive Medicine and Biostatistics, Uniformed Services University, Bethesda, MD; Department of Medicine, Uniformed Services University of the Health Sciences, Bethesda, MD; Brooke Army Medical Center, Joint Base San Antonio-Ft Sam Houston, TX; Departments of Internal Medicine and Anesthesiology, Wake Forest School of Medicine, Winston-Salem, North Carolina; Naval Medical Center, San Diego, California; Fort Belvoir Community Hospital, Fort Belvoir, VA; Madigan Army Medical Center, Joint Base Lewis-McChord, WA; Walter Reed National Military Medical Center, Bethesda, MD; Naval Medical Center Portsmouth, Portsmouth, VA; Department of Pediatrics, Uniformed Services University, Bethesda, MD; Department of Pharmacology & Molecular Therapeutics, Uniformed Services University, Bethesda, MD; Biological Defense Research Directorate, Naval Medical Research Center-Frederick, Ft. Detrick, MD

**Keywords:** COVID-19, SARS-CoV-2, Topological Data Analysis, Coronavirus Infections, immunology, Cytokines, analysis

## Abstract

**OBJECTIVES:** The relationships between baseline clinical phenotypes and the cytokine milieu of the peak ‘inflammatory’ phase of coronavirus 2019 (COVID-19) are not yet well understood. We used Topological Data Analysis (TDA), a dimensionality reduction technique to identify patterns of inflammation associated with COVID-19 severity and clinical characteristics.

**DESIGN:** Exploratory analysis from a multi-center prospective cohort study.

**SETTING:** Eight military hospitals across the United States between April 2020 and January 2021.

**PATIENTS:** Adult (≥18 years of age) SARS-CoV-2 positive inpatient and outpatient participants were enrolled with plasma samples selected from the putative ‘inflammatory’ phase of COVID-19, defined as 15-28 days post symptom onset.

**INTERVENTIONS:** None.

**MEASUREMENTS AND MAIN RESULTS:** Concentrations of 12 inflammatory protein biomarkers were measured using a broad dynamic range immunoassay. TDA identified 3 distinct inflammatory protein expression clusters. Peak severity (outpatient, hospitalized, ICU admission or death), Charlson Comorbidity Index (CCI), and body mass index (BMI) were evaluated with logistic regression for associations with each cluster. The study population (n=129, 33.3% female, median 41.3 years of age) included 77 outpatient, 31 inpatient, 16 ICU-level, and 5 fatal cases. Three distinct clusters were found that differed by peak disease severity (p <0.001), age (p <0.001), BMI (p<0.001), and CCI (p=0.001).

**CONCLUSIONS:** Exploratory clustering methods can stratify heterogeneous patient populations and identify distinct inflammation patterns associated with comorbid disease, obesity, and severe illness due to COVID-19.

## Background

While clinical risk factors for coronavirus disease 2019 (COVID-19) severity have been described, mechanisms of inflammation associated with these baseline clinical features are less understood (1). SARS-CoV-2 infections range from asymptomatic to fatal illness. This spectrum is associated with host risk factors such as age and chronic noncommunicable disease (NCD), including obesity and cardiovascular disease (2). However, the pathways from host factors to COVID-19 severity and sequelae are largely unknown. Given the heterogeneity of COVID-19 severity and a growing immunomodulatory treatment armamentarium (2, 3), pathologic inflammation patterns and their association with comorbidities need to be identified to optimize treatment selection.

COVID-19 severity and inflammation occur in three phases: acute, inflammatory, and late phases of illness. Peak severity and peak inflammatory biomarkers generally occur after two weeks of illness (15 to 28 days after symptom onset) during the inflammatory phase (4, 5). While inflammation may subside in mild cases, persistently high proinflammatory cytokines have been noted in more severe cases during this period. This time window of heightened immune response may be best suited to elucidate the relationship between host factors and severe COVID-19. *In silico* stratification of host-biomarker profiles using exploratory clustering and machine learning analyses has the potential to identify distinct phenotypes associated with disease severity, which in turn can lead to discovery of personalized treatment approaches.

Herein we define inflammatory host-biomarker phenotypes of COVID-19 identified by Topological Data Analysis (TDA) and their associated comorbid conditions and disease severity. TDA is a multivariate pattern analytical tool that uses an unsupervised approach to dimensionality reduction and data visualization (6). TDA can be used to identify biomarker patterns and phenotype-biomarker relationships (7-9). TDA has been demonstrated to identify patient subgroups that would benefit from personalized interventions for heterogenous diseases such as cancer care and primary ciliary dyskinesia (6, 8). We hypothesized that the network approach of TDA clustering would identify unique inflammation phenotype patterns associated with severity, demographics, and co-morbid conditions known to predispose patients to worse outcomes during SARS-CoV-2 infection (4). Our analysis focused on samples collected during the inflammatory phase from an observational cohort of participants with mild to severe COVID-19 at military treatment facilities. Inflammatory biomarkers were selected from prior unpublished non-COVID-19 sepsis TDA analyses (10) and from clinical use (11). We sought to demonstrate that this analytical approach can help discern inflammatory patterns to find possible treatment targets, as well as serve as a tool to understand baseline host factors and severe COVID-19.

## Methods

Participants were enrolled in a prospective, multi-center COVID-19 cohort under the Epidemiology, Immunology, and Clinical Characteristics of Emerging Infectious Diseases with Pandemic Potential (EPICC) protocol, at 8 military treatment facilities (Brooke Army Medical Center, San Antonio, TX; Fort Belvoir Community Hospital, Fort Belvoir, VA; Madigan Army Medical Center, Joint Base Lewis-McChord, WA; Naval Medical Center Portsmouth, Portsmouth, VA; Naval Medical Center San Diego, San Diego, CA; Tripler Army Medical Center, Honolulu, HI; William Beaumont Army Medical Center, El Paso, TX; Walter Reed National Military Medical Center, Bethesda, MD) between April 2020 and January 2021 (12). The protocol was approved by the Uniformed Services University Institutional Review Board (IDCRP-085)(13). All patients provided written informed consent. EPICC study enrollment included subjects ≥18 years of age with laboratory-confirmed or suspected SARS-CoV-2 infection seeking inpatient or outpatient medical care. Following consent, demographic, comorbidity, and illness data were collected through participant interviews and a review of the participant’s electronic medical record or using participant completed surveys implemented in November 2020. Subjects with a positive clinical SARS-CoV-2 RT-PCR result and plasma samples collected were included in this analysis. Results of well-described (14) COVID-19 clinical biomarkers CRP, ferritin, and IL-6, were explored from 249 participants with plasma collected 0-29 days post symptom onset (dpso) to determine if the longitudinal inflammatory biomarker LOESS (locally estimated scatterplot smoothing) curve peaked between 14 to 28 days per previously published phases of illness framework for studying COVID-19 (Supplementary Figure S1) (4). These trends were consistent with the literature, and, accordingly, the TDA was restricted to the 129 participants with samples collected during the inflammatory phase defined as 15-28 dpso. Receipt of baricitinib, tocilizumab, hydroxychloroquine, or systemic steroids (equivalent to prednisone 10mg daily or above) at the time of blood collection was determined through the electronic medical record or participant surveys.

Plasma samples were prospectively collected after enrollment as previously described (13). Venous whole blood samples were centrifuged for 10 minutes at 1500 g and collected plasma was stored at −80°C. A panel of 12 inflammatory proteins were measured in the plasma samples using the high dynamic range automated enzyme-linked immunosorbent assay Ella microfluidic analyzer (ProteinSimple, San Jose, California, USA). The panel included: IL-6, CXCL10, IL-1RA, D-dimer, procalcitonin, ferritin, VEGF-A, IL-5, soluble receptor for advanced glycation end-product (RAGE), TNFR1, IFN-γ, and C-reactive protein (CRP). This panel was selected to include analytes in clinical use for prognostication (i.e., CRP, procalcitonin, ferritin, and D-dimer)(11), based on prior COVID-19 literature (i.e., IL-6, IFN-γ and CXCL10) (15), and identified to be representative of prior TDA-based non-COVID-19 sepsis clusters (i.e., IL-1RA, VEGF-A, IL-5, RAGE, and TNFR1) (10, 16). All protein concentrations were log_10_-transformed and normalized for site-to-site variation using the *R* package *SVA ComBat* (17). A small number (1.6%) of missing values were imputed using a *k*-nearest neighbor model, and out-of-range values were imputed using either the lowest or highest measured value within range of the Ella platform. Correlation between analytes was explored with a principal component analysis and determining the Spearman’s correlation coefficients. For some subjects, multiple samples were available. In such cases, the sample with the highest coefficient of variation across all analytes was retained to incorporate the largest degree of relative variability at that time point (18).

Protein expression networks were generated solely using biomarkers levels with the TDA “Mapper” algorithm using the EurekaAI platform (SymphonyAI, Los Altos, CA, USA)(7, 19, 20). TDA networks were generated for a range of resolution settings to examine the persistence of subject clusters and their interrelatedness. Peaked severity (outpatient, hospitalized, ICU-level or death) color gradients were overlaid on identified clusters. Levels of the individual proteins in each TDA group were summarized in a series of boxplots (*R* package “ggplot2” v3.3.5). Backward selection stepwise logistic regression using a Bernoulli-adjusted significance level of 0.0042 (i.e., 0.05/12) was used to identify which proteins were up- or downregulated within each cluster. While TDA clusters will inherently have different biomarker levels, this was performed to simplify inference about representative biomarkers and for future validation in external cohorts. A sensitivity analysis was performed adjusting for peak severity to determine the effect of covariate selection. An additional sensitivity analysis was performed excluding participants receiving systemic steroids.

Summary statistics were calculated for the TDA clusters, comparing baseline demographics (e.g., sex, age, race, ethnicity, selected medical comorbidities), days post symptom onset, peak severity, steroid use, and the inflammatory biomarkers by clusters using either Chi-square (categorical values), Fisher exact (categorical values), or Mann-Whitney U tests (continuous values). Charlson Comorbidity Index (CCI) and body mass index (BMI) values were divided into score-based categories (i.e., CCI: 0, 1-2, 3-4, or 5+; BMI: <30, 30-39.9, or ≥40 kg/m^2^) to describe the prevalence of comorbid conditions by cluster on a bar plot but were otherwise treated as continuous values. BMI values were not available from 6.2% of the cohort. Peak severity was categorized for each participant (outpatient, non-ICU [intensive care unit] inpatient, and ICU or death). Multivariable logistic regression adjusting for peak severity was used to identify associations between each TDA cluster and BMI or CCI at a significance level of 0.05. All statistical analyses were performed in Stata (version 15.0; StataCorp LLC, College Station, TX, USA) and R version 4.0.2 (21)

## Results

Biomarkers CRP, IL-6, and ferritin were stratified by severity and explored for the 249 participants in the EPICC cohort between 0-28 dpso using a scatter plot with LOESS (locally estimated scatterplot smoothing) curves. This demonstrated average cytokines peaked or remained elevated during the described inflammatory phase (15-28 dpso) among ICU-level or fatal courses of illness (Supplementary Figure S1). Based on these findings and the inflammatory phase literature, we restricted our analysis to 129 participants (66.7% male, median 41.3 years of age) including 77 outpatient, 31 inpatient, 16 ICU-level, and 5 fatal cases (Table 1) between 15 to 28 days of illness. Correlation along a PCA axis was observed among procalcitonin, TNFR1, IL-6, CRP, and IL-1RA while RAGE, IFN-γ, IL-5, and VEGF-A were less correlated with the other analytes. Additionally, variance increased with each level of peak severity (Supplementary Figure S2). These results supported the additive information provided by the 12 protein analytes, and TDA was performed. Interestingly, 3 distinct inflammatory proteins clusters, labeled Cluster 1, Cluster 2, and Cluster 3 (Figure 1; Supplementary Figure S3), were consistently identified using TDA.

**Table 1.**
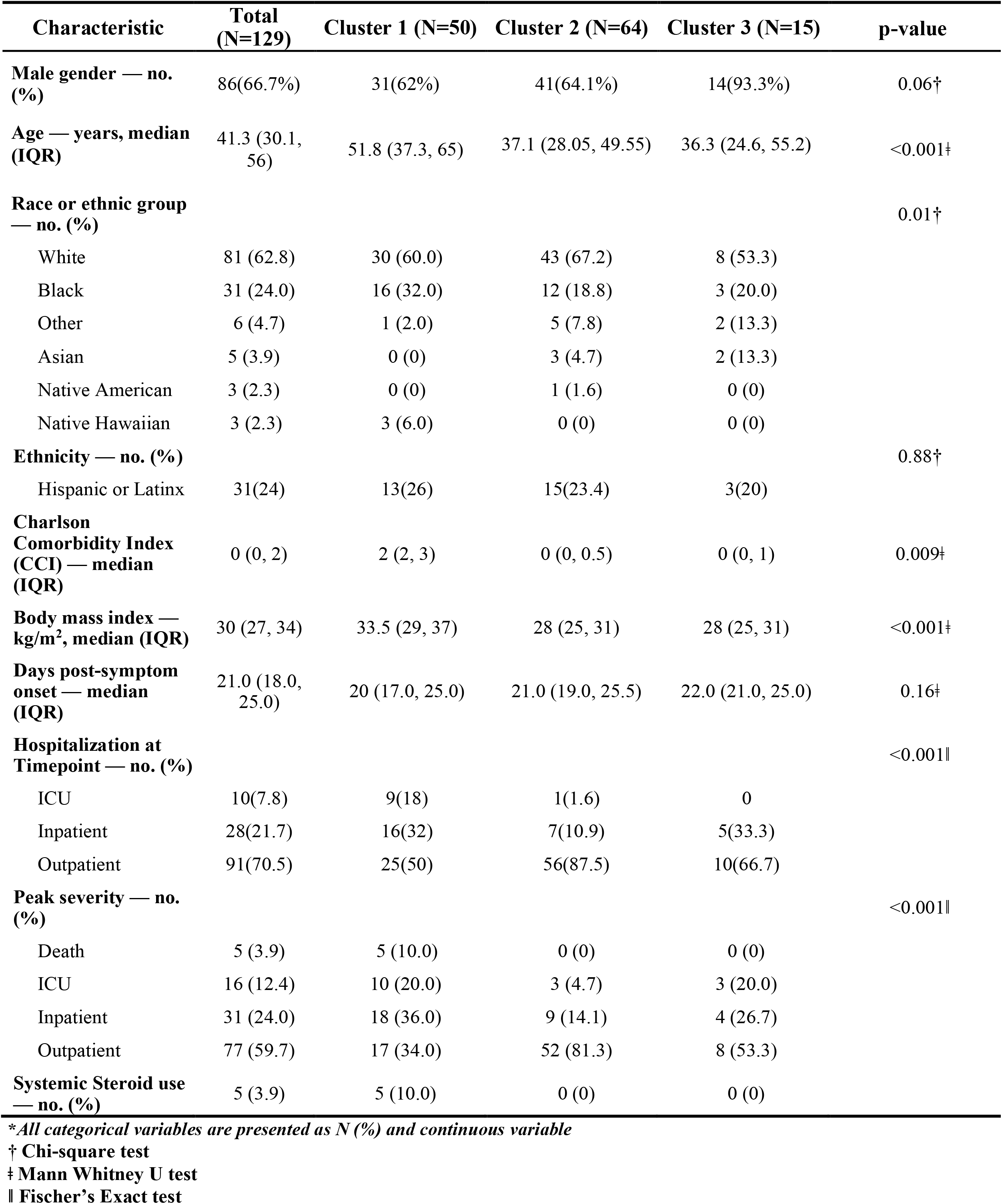
Baseline demographics across TDA clusters.

**Figure 1.**
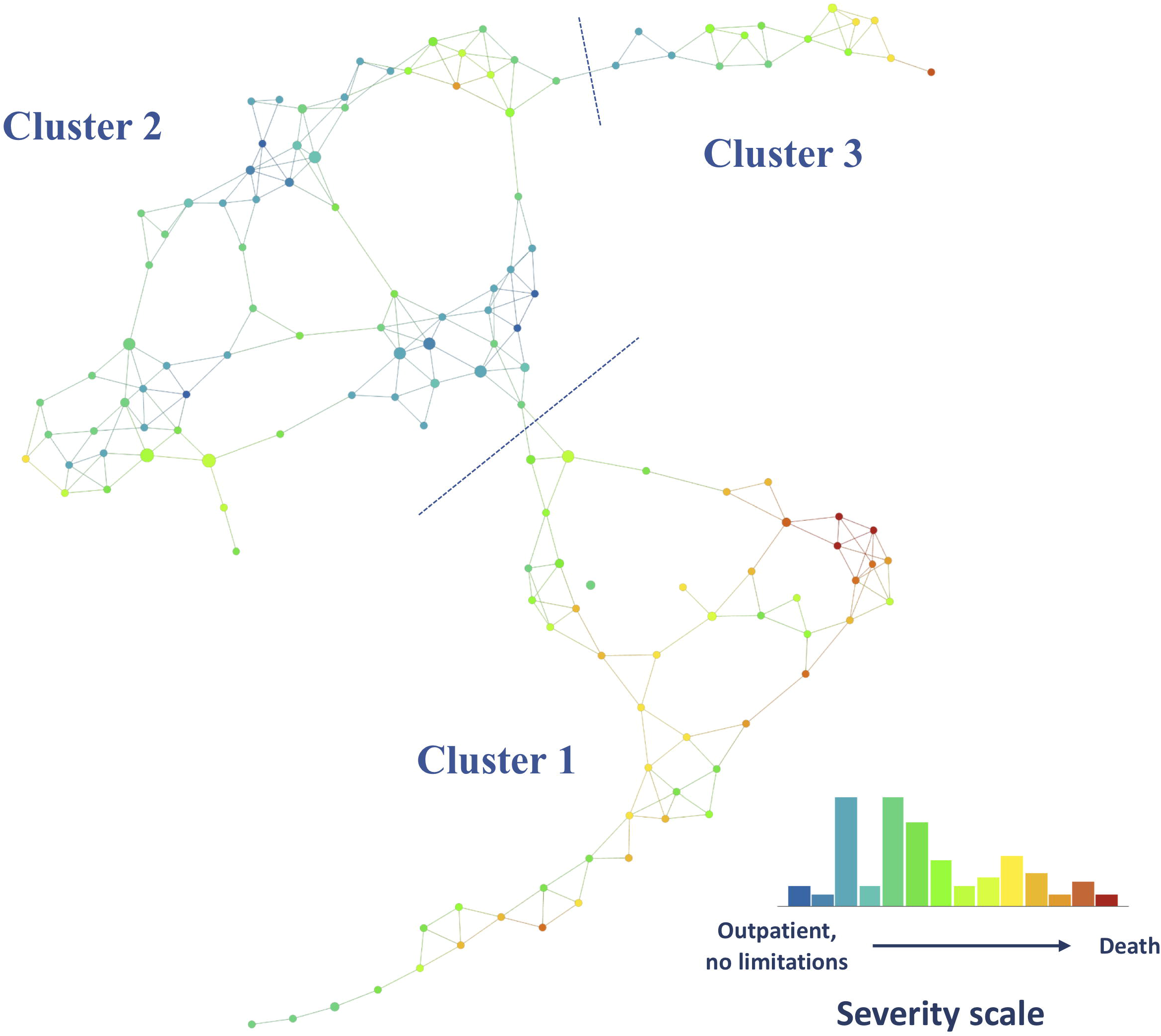
Topological data analysis (TDA) network of protein expression during the middle-phase of COVID-19. Distinct protein expression phenotypes (Clusters 1, 2, and 3) were identified based on density and break points in the network and persistence of the clusters. Each node represents a combination of 12 plasma protein analyte levels and its size increases with the number of participants that are included. Edges (lines between nodes) indicate that patients are represented in more than one node. The network is colored by the average score on the disease severity scale (from outpatients without limitations [green] to death [red]) in each node. Analysis was performed on the EurekaAI Workbench (SymphonyAI, Los Altos, CA, USA).

Age differed significantly between TDA clusters (p<0.001). Participants from TDA Clusters 2 (median 37.1 years of age; IQR, 28.1 to 50.0) and 3 (median 36.3 years of age; IQR, 24.6 to 55.2) were younger than in Cluster 1 (median 51.8 years of age; IQR, 37.3 to 65.0) (Table 1). The prevalence of male gender was similar among Cluster 1 (62.0%, n=31), Cluster 2 (64.1%, n=41), and the general cohort (66.7%), but cluster 3 was predominantly male (93.3%, n=14). The median time from symptom onset to sample collection was 21 days (IQR 18 to 25) and did not differ between clusters (Table 1). Peak disease severity, as categorized by hospitalization status, was also found to differ significantly among the TDA clusters (p<0.001). Cluster 1 had the highest prevalence of severe COVID-19, comprising 66.0% (n=33) hospitalized participants, compared to 46.7% (n=7) hospitalized participants in Cluster 3, and 18.8% (n=12) hospitalized participants in Cluster 2 (Figure 2, Table 1). All fatal cases (n=5) were in Cluster 1. No individuals had received baricitinib or tocilizumab, and hydroxychloroquine use was limited to 2 individuals in Cluster 1. Receipt of systemic steroids at the time of blood collection was limited to 5 participants in Cluster 1 (10.0%; n=5).

**Figure 2.**
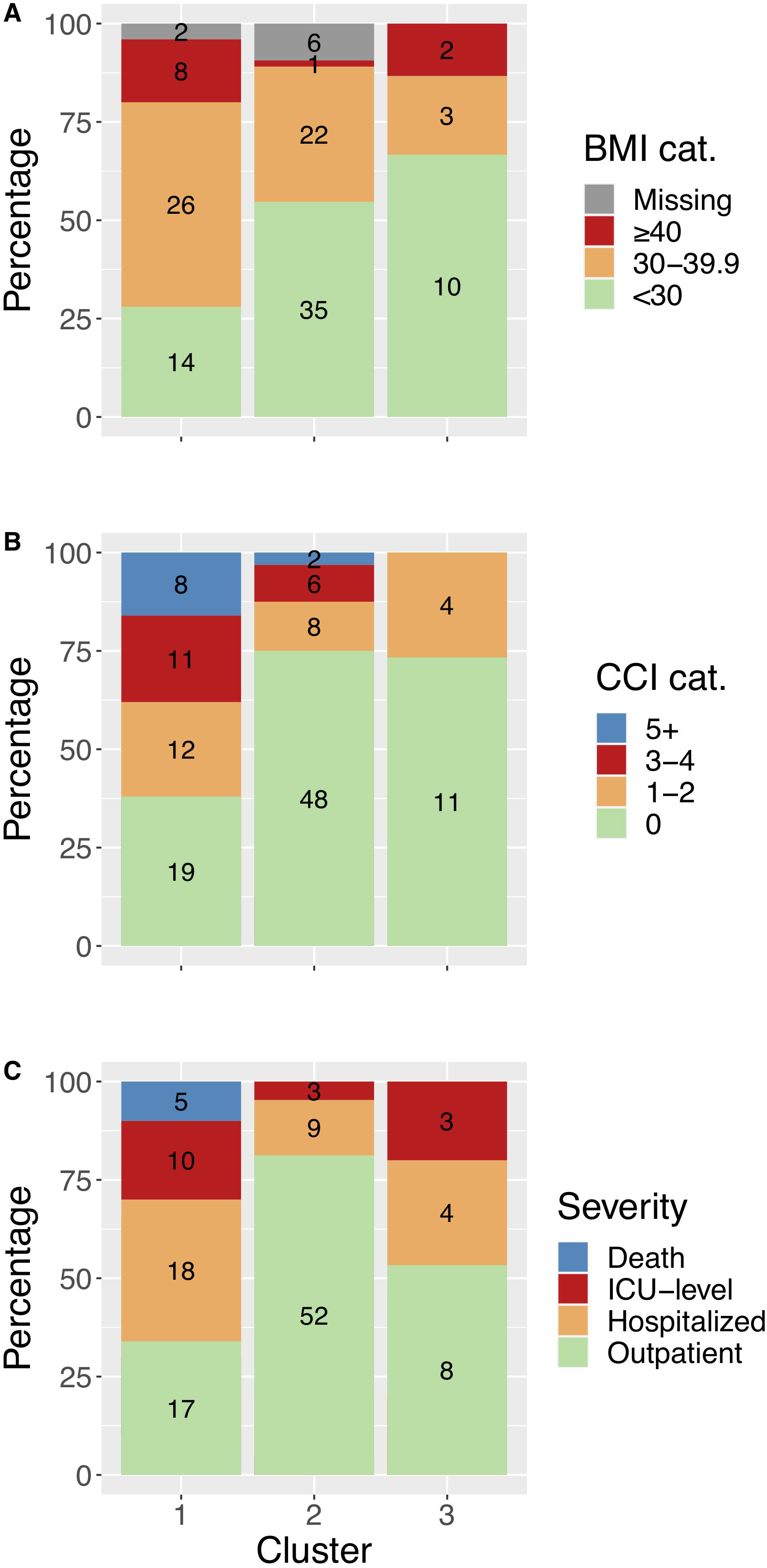
Cluster differences with bar plots (% [n]) of comorbid diseases and severity by cluster. A: BMI (body mass index) category (range in kg/m^2^) prevalence by cluster; B: Charlson Comorbidity Index (CCI) category prevalence by cluster; C: peak levels of severity by cluster. Total (n) presented in the center of each category.

The median CCI differed (p=0.009) among clusters ranging from 2 (IQR, 2 to 3) in Cluster 1 to 0 (IQR 0 to 0.5) in Cluster 2 and 0 (IQR, 0 to 1) in Cluster 3. Most participants in Cluster 2 (75.0%) and in Cluster 3 (73.3%) had a CCI of 0 compared to 38.0% of individuals in Cluster 1 (Figure 2). Additionally, median BMI was higher in Cluster 1 (33.5 kg/m^2^; IQR, 29.0 to 37.0) compared to in Cluster 2 and Cluster 3, which were the same (28.0 kg/m^2^; IQR, 25.0 to 31.0)(Table 1). After adjusting for peak severity using logistic regression, participants with a higher BMI (OR: 1.1 per kg/m^2^, p=0.002) and a higher CCI (OR: 1.3 for each score increase, p=0.02) were more common in Cluster 1 compared to participants in Cluster 2 and 3 combined.

In summary, participants in Cluster 1 were more likely to be older, have higher BMI and more comorbidities, and have more severe disease, whereas participants in Cluster 2 were more likely to be younger, have lower BMI and comorbidities, and have mild illness (Figure 2). Cluster 3 was predominantly composed of younger adult predominantly male participants, without comorbid conditions, among whom almost half (7 of 15) were hospitalized.

The distributions of each analyte were different across clusters using a chi-squared test, except for IL-5 and IFN-γ which had a similar distribution (Table 2). Certain biomarkers including CRP, IL-6, IL-1RA, D-dimer, TNFR1, and VEGF-A were more elevated in Cluster 1 (older participants with higher severity) compared to Clusters 2 and 3 (Table 2; Figure 3; Supplementary Figure 4). RAGE was lower in Cluster 1 compared to Clusters 2 or 3 and IFN-γ was lower in Cluster 1 compared to Cluster 2 (Figure 3; Supplementary Figure 4). Cluster 3, a young cluster with moderate severity, was found to have higher ferritin, procalcitonin, and CXCL10, and lower VEGF-A compared to Cluster 2, a similarly young cluster with mild illness.

**Table 2.**
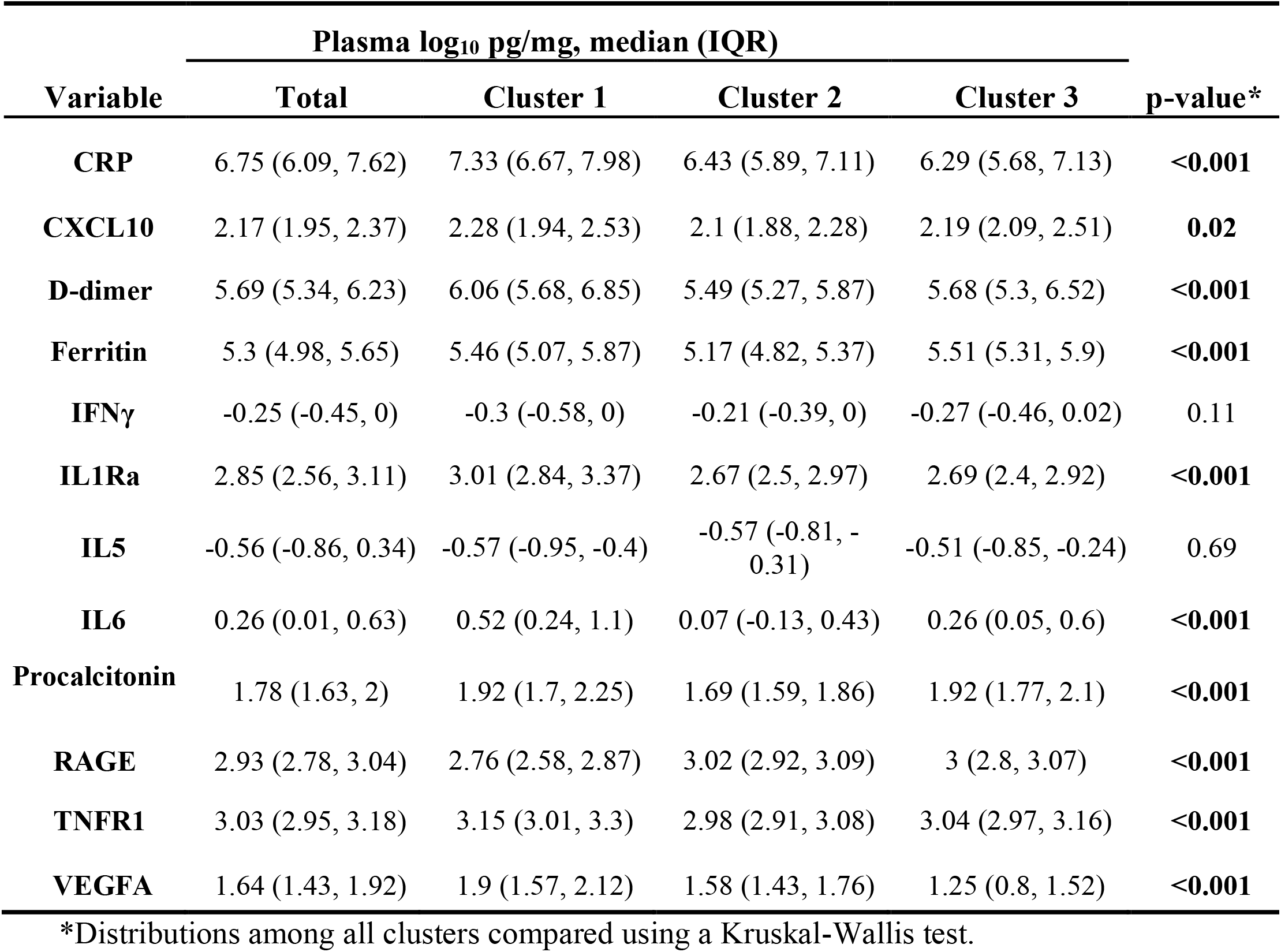
Comparison of the Ella biomarkers across TDA clusters. For each subject, one sample was selected based on highest coefficient of variation.

| Variable | Plasma log<sub>10</sub> pg/mg, median (IQR) |  |  |  | p-value* |
| --- | --- | --- | --- | --- | --- |
|  | Total | Cluster 1 | Cluster 2 | Cluster 3 |  |
| **CRP** | 6.75 (6.09, 7.62) | 7.33 (6.67, 7.98) | 6.43 (5.89, 7.11) | 6.29 (5.68, 7.13) | **<0.001** |
| **CXCL10** | 2.17 (1.95, 2.37) | 2.28 (1.94, 2.53) | 2.1 (1.88, 2.28) | 2.19 (2.09, 2.51) | **0.02** |
| **D-dimer** | 5.69 (5.34, 6.23) | 6.06 (5.68, 6.85) | 5.49 (5.27, 5.87) | 5.68 (5.3, 6.52) | **<0.001** |
| **Ferritin** | 5.3 (4.98, 5.65) | 5.46 (5.07, 5.87) | 5.17 (4.82, 5.37) | 5.51 (5.31, 5.9) | **<0.001** |
| **IFNγ** | -0.25 (-0.45, 0) | -0.3 (-0.58, 0) | -0.21 (-0.39, 0) | -0.27 (-0.46, 0.02) | 0.11 |
| **IL1Ra** | 2.85 (2.56, 3.11) | 3.01 (2.84, 3.37) | 2.67 (2.5, 2.97) | 2.69 (2.4, 2.92) | **<0.001** |
| **IL5** | -0.56 (-0.86, 0.34) | -0.57 (-0.95, -0.4) | -0.57 (-0.81, -0.31) | -0.51 (-0.85, -0.24) | 0.69 |
| **IL6** | 0.26 (0.01, 0.63) | 0.52 (0.24, 1.1) | 0.07 (-0.13, 0.43) | 0.26 (0.05, 0.6) | **<0.001** |
| **Procalcitonin** | 1.78 (1.63, 2) | 1.92 (1.7, 2.25) | 1.69 (1.59, 1.86) | 1.92 (1.77, 2.1) | **<0.001** |
| **RAGE** | 2.93 (2.78, 3.04) | 2.76 (2.58, 2.87) | 3.02 (2.92, 3.09) | 3 (2.8, 3.07) | **<0.001** |
| **TNFR1** | 3.03 (2.95, 3.18) | 3.15 (3.01, 3.3) | 2.98 (2.91, 3.08) | 3.04 (2.97, 3.16) | **<0.001** |
| **VEGFA** | 1.64 (1.43, 1.92) | 1.9 (1.57, 2.12) | 1.58 (1.43, 1.76) | 1.25 (0.8, 1.52) | **<0.001** |

\*Distributions among all clusters compared using a Kruskal-Wallis test.

**Figure 3.**
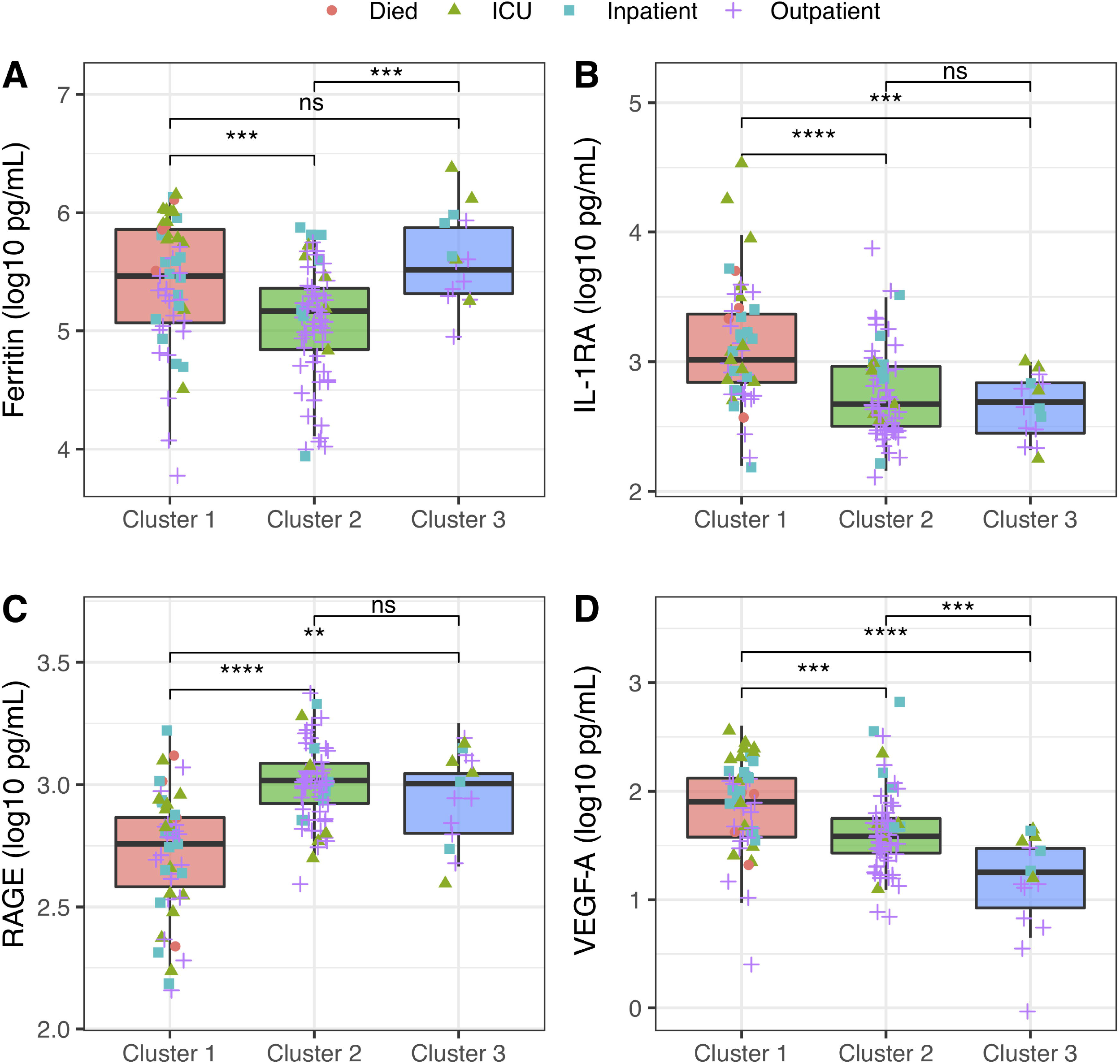
Box plots of markers selected in stepwise regression to identify characteristic biomarkers of each cluster: Ferritin (A), IL1RA (B), RAGE (C), and VEGFA (D) by cluster. Kruskal-Wallis test performed comparing analyte levels between clusters. **: p ≤ 0.01; ***: p ≤ 0.001; ****: p ≤ 0.0001

Stepwise regression, both unadjusted and adjusted for peak severity, was used to identify which analytes were most characteristic of each TDA cluster (Supplementary Table S1.). The distinguishing biomarker of Cluster 1 were relatively high IL-1RA and low RAGE levels; these subjects had a high severity phenotype compared to other clusters (Figure 3; Supplementary Figure 4; Supplementary Table S1.). Regardless of peak severity, Cluster 2 was characterized by relatively low procalcitonin and high RAGE levels. Cluster 3 was characterized by low VEGF-A after peak severity adjustment (Figure 3; Supplementary Figure 4; Supplementary Table S1.). When restricting the analysis to those not receiving steroids, the models were qualitatively unchanged, and the same covariates were selected.

## Discussion

We demonstrated that a multi-site prospective patient cohort can be stratified into three distinct inflammatory profiles using 12 protein biomarkers from samples collected during the inflammatory phase of COVID-19. TDA dimensionality reduction was able to identify biomarker patterns with differences in both severity and comorbid conditions between cluster phenotypes. Combinations of biomarkers, independent of clinical information, grouped participants into one of three distinct clusters: high COVID-19 severity, older, with comorbid conditions (Cluster 1); low severity, younger, less comorbid illness (Cluster 2); and a moderate severity, younger, previously healthy, male-predominant group (Cluster 3). This proof-of-concept study identifies potential use of TDA as a strategy to identify biomarker clusters associated with the heterogeneity of COVID-19 clinical presentations. Whilst exploratory, this reveals potential translational approaches to using host-biomarker stratification with advanced clustering and network analytical techniques, such as TDA, to better understand what drives phenotypic differences in the clinical presentation of COVID-19.

Patterns of inflammation observed for the different TDA clusters could suggest dysregulated pathways associated with COVID-19 pathology. Cluster 1 was found to be the highest severity cluster with all fatal cases and most ICU-level cases. This cluster contained distinctly more subjects with baseline comorbid conditions and obesity as defined by BMI ≥30. Cluster 1 subjects had higher IL-1RA compared to Cluster 2 and 3, clusters represented by participants with less comorbid conditions. Consistent with this trend, prior work has identified IL-1RA as a potential mediator between obesity and COVID-19 severity (22). Interestingly, IFN-γ was lower and IL-6 higher in Cluster 1 compared to the Cluster 2 participants. This pattern of an aberrant Th1 response has been previously identified to be associated with severe COVID-19 and potentially distinct from influenza infection (22). Cluster 1 aligned with baseline comorbid illnesses known to be risk factors for severe COVID-19 with potentially distinct inflammatory cascade patterns demonstrated.

Cluster 3 was unique in that it had a combination of low VEGF-A but had elevated ferritin and higher prevalence of severe illness compared to Cluster 2, a mild illness cluster with comparable demographics. While sample size is limited, 14 of 15 participants in Cluster 3 were male, suggestive of a biologic sex difference in immune response among these previously healthy young men. Sex differences leading to severe COVID-19 among men have been previously described with X-linked TLR7 deficiency(23, 24) and on a larger scale with sex-related differences in innate and T-cell responses (25). A combination of low VEGF-A and elevated ferritin may identify a unique inflammation subtype and merits further study with external cohorts.

RAGE, a biomarker of acute lung injury (26), was found to have different distributions between clusters. In contrast to prior research (27), RAGE levels appeared to be higher among the younger and relatively milder COVID-19 severity Cluster 2 compared to Cluster 1. Compared to other clusters, RAGE was elevated along with IFN-γ in the less symptomatic Cluster 2, but with lower acute phase reactants ferritin and procalcitonin. The converse was true with Cluster 1 where lower levels of RAGE in individuals were noted, along with elevated acute phase reactants (i.e., CRP, procalcitonin, and ferritin). This association of lower RAGE with higher severity Clusters 1 and 3 contrasts with a direct association with COVID-19 mortality (28). However, our results may differ by accounting for biomarker patterns rather than evaluating each biomarker in isolation. It is possible that RAGE could be an indicator of severity during certain disease states but functioning as an adaptive anti-inflammatory protein in Cluster 2 during the inflammatory phase. Soluble RAGE has been shown to reduce vascular injury in rodent models (29, 30) and could be protective against vascular inflammation mediated the RAGE receptor (31). The paradoxically inverse relationship observed between RAGE and these commonly used acute phase reactants between the clusters could be useful for identification and stratification of individuals with COVID-19.

While this study, to our knowledge, is the first to use an advanced dimensionality reduction approach to understand relationships between biomarker patterns and clinical phenotypes during the inflammatory phase of COVID-19, there are limitations worth noting. Samples were collected from April 2020 to January 2021 and treatment practices and epidemiologic changes over time may have affected inflammation patterns. Hence, we incorporated a sensitivity analysis excluding those that received systemic steroids in Cluster 1 to aid in interpreting the findings. In addition, the sample size may limit our ability to identify uncommon biomarker patterns and external validation is needed of patterns identified. Additionally, regression was used to adjust for peak severity to identify biomarker and comorbid condition associations with TDA clusters distinct from severity trajectory differences. While this is a novel feature of this biomarker study, residual confounding related to peak severity remains possible. Despite limitations, results presented here are hypothesis generating and should be evaluated further in additional cohorts.

This approach constitutes an early exploratory step in identifying host biomarker patterns that may be leveraged for personalized interventions, and offers new insights for COVID19 prognosis, therapy, and prevention with techniques that could be extended to understanding other severe infections. Using analytes identified from our international sepsis cohort research(10), 3 biomarker clusters with different phenotypic associations were identified among those with heterogenous COVID-19 presentations. The application of these biomarkers derived from non-COVID-19 severe infection research suggests that pathogen-agnostic sepsis biomarkers could be identified for personalized approaches to triage of care or immunomodulation strategies. Further validation of these markers and clustering algorithms with external cohorts could inform point-of-care biomarker assay development to guide more individualized approaches to COVID-19 care.

## Data Availability

All data produced in the present study are available upon reasonable request to the authors

## Acknowledgements

**We thank the members of the EPICC COVID-19 Cohort Study Group for their many contributions in conducting the study and ensuring effective protocol operations. The following members were all closely involved with the design, implementation, and/or oversight of the study and have met group authorship criteria for this manuscript:

*Brooke Army Medical Center, Fort Sam Houston, TX:* Col J. Cowden; LTC M. Darling; T. Merritt; CPT

T. Wellington

*Fort Belvoir Community Hospital, Fort Belvoir, VA:* A. Rutt

*Madigan Army Medical Center, Joint Base Lewis McChord, WA:* CAPT C. Conlon; COL P. Faestel; COL C. Mount

*Naval Medical Center Portsmouth, Portsmouth, VA:* LCDR A. Smith; R. Tant; T. Warkentien *Naval Medical Center San Diego, San Diego, CA:* CDR C. Berjohn; CAPT (Ret) G. Utz *Tripler Army Medical Center, Honolulu, HI:* LTC C. Madar; C. Uyehara

*Uniformed Services University of the Health Sciences, Bethesda, MD:* K Chung; C. English; C. Fox; M.

Grother; COL P. Hickey; E. Laing; LTC J. Livezey; E. Parmelee; J. Rozman; M. Sanchez; A. Scher

*United States Air Force School of Aerospace Medicine, Dayton, OH:* Sgt T. Chao; R. Chapleau; A. Fries;

K. Reynolds

*Womack Army Medical Center, Fort Bragg, NC:* LTC D. Hostler; LTC J. Hostler; MAJ K. Lago; C. Maldonado

*William Beaumont Army Medical Center, El Paso, TX:* MAJ T. Hunter; R. Mody; M. Wayman

*Walter Reed National Military Medical Center, Bethesda, MD:* MAJ N. Huprikar

The authors wish to also acknowledge all who have contributed to the EPICC COVID-19 study:

*Brooke Army Medical Center, Fort Sam Houston, TX:* Col J. Cowden; LTC M. Darling; S.

DeLeon; Maj D. Lindholm; LTC A. Markelz; K. Mende; S. Merritt; T. Merritt; LTC N. Turner; CPT T. Wellington

*Carl R. Darnall Army Medical Center, Fort Hood, TX:* LTC S. Bazan; P.K Love

*Fort Belvoir Community Hospital, Fort Belvoir, VA:* N. Dimascio-Johnson; MAJ E. Ewers; LCDR K. Gallagher; LCDR D. Larson; A. Rutt

*Henry M. Jackson Foundation, Inc*., *Bethesda, MD:* P. Blair; J. Chenoweth; D. Clark

*Madigan Army Medical Center, Joint Base Lewis McChord, WA:* S. Chambers; LTC C. J. Colombo; R. Colombo; CAPT C. Conlon; CAPT K. Everson; COL P. Faestel; COL T. Ferguson; MAJ L. Gordon; LTC S. Grogan; CAPT S. Lis; COL C. Mount; LTC D. Musfeldt; CPT D. Odineal; LTC M. Perreault; W. Robb-McGrath; MAJ R. Sainato; C. Schofield; COL C. Skinner; M. Stein; MAJ M. Switzer; MAJ M. Timlin; MAJ S. Wood

*Naval Medical Center Portsmouth, Portsmouth, VA:* S. Banks; R. Carpenter; L. Kim; CAPT

K. Kronmann; T. Lalani; LCDR T. Lee; LCDR A. Smith; R. Smith; R. Tant; T. Warkentien *Naval Medical Center San Diego, San Diego, CA:* CDR C. Berjohn; S. Cammarata; N. Kirkland; CAPT (Ret) R. Maves; CAPT (Ret) G. Utz

*Tripler Army Medical Center, Honolulu, HI:* S. Chi; LTC R. Flanagan; MAJ M. Jones; C. Lucas; LTC C. Madar; K. Miyasato; C. Uyehara

*Uniformed Services University of the Health Sciences, Bethesda, MD:* B. Agan; L. Andronescu; A. Austin; C. Broder; CAPT T. Burgess; C. Byrne; COL K Chung; J. Davies; C. English; N. Epsi; C. Fox; M. Fritschlanski; M. Grother; A. Hadley; COL P. Hickey; E. Laing; LTC C. Lanteri; LTC

J. Livezey; A. Malloy; R. Mohammed; C. Morales; P. Nwachukwu; C. Olsen; E. Parmelee; S. Pollett; S.

Richard; J. Rozman; J. Rusiecki; E. Samuels; M. Sanchez; A. Scher; CDR M. Simons; A. Snow; K. Telu; D. Tribble; L. Ulomi

*United States Air Force School of Medicine, Dayton, OH:* Sgt T. Chao; R. Chapleau; A. Fries; C. Harrington; S. Huntsberger; S. Purves; K. Reynolds; J. Rodriguez; C. Starr

*Womack Army Medical Center, Fort Bragg, NC:* B. Barton; LTC D. Hostler; LTC (Ret) J. Hostler; MAJ

K. Lago; C. Maldonado; J. Mehrer

*William Beaumont Army Medical Center, El Paso, TX:* MAJ T. Hunter; J. Mejia; R. Mody; R. Resendez; P. Sandoval; M. Wayman

*Walter Reed National Military Medical Center, Bethesda, MD:* I. Barahona; A. Baya; A. Ganesan; MAJ N. Huprikar; B. Johnson

*Walter Reed Army Institute of Research, Silver Spring, MD:* S. Peel

## Disclaimer

The contents of this article are the sole responsibility of the authors and do not necessarily reflect the views, assertions, opinions, or policies of the Henry M. Jackson Foundation for the Advancement of Military Medicine, Inc., the U.S. Department of Defense, the U.S. government, or any other government or agency. Mention of trade names, commercial products, or organizations does not imply endorsement by the U.S. government. Some of the authors of this work are military service members or employees of the U.S. government. This work was prepared as part of their official duties. Title 17 U.S.C. x105 provides that ‘‘Copyright protection under this title is not available for any work of the United States government.’’ Title 17 U.S.C. x101 defines a U.S. government work as a work prepared by a military service member or employee of the U.S. government as part of that person’s official duties. The investigators have adhered to the policies for protection of human subjects as prescribed in 45 CFR 46. This research has been approved the USU Institutional Review Board in compliance with all applicable federal regulations governing the protection of human subjects.

