## Supplemental figures and tables. for "Topological data analysis identifies distinct biomarker phenotypes during the ‘inflammatory’ phase of COVID-19"

**Table S1.** Multivariable logistic regression models for clusters and plasma immune biomarker levels. Models were fit comparing each cluster against all other clusters.

| Cluster | Covariates in unadjusted model | OR* (95% CI) | AIC | Covariates in severity-adjusted** model | OR* (95% CI) | AIC |
|---|---|---|---|---|---|---|
| **1** | | | 87.7 | | | 85.4 |
| | CXCL10 | 238.1 (6.1, 9,238.2) | | Severity | 4.9 (2.0, 11.9) | |
| | IL1RA | 4.3 (2.1, 8.9) | | IL1RA | 4.1 (2.1, 8.2) | |
| | RAGE | 0.0007 (0.00005, 0.01) | | RAGE | 0.001 (0.00008 0.02) | |
| **2** | | | 91.9 | | | 102.0 |
| | Ferritin | 1.0 (0.990, 0.997) | | Severity | 0.4 (0.1, 0.9) | |
| | Procalcitonin | 2.5e-14(1.7e-22, 3.6e-06) | | Procalcitonin | 2.8e-14 (06.5e-22, 1.2e-06) | |
| | RAGE | 839.1 (58.7, 11,991.9) | | RAGE | 217.0 (24.2, 1,948.7) | |
| **3** | | | 58.7 | | | 73.4 |
| | Ferritin | 1.0 (1.002, 1.007) | | Severity | 1.9 (0.9, 3.9) | |
| | VEGFA | 3.1e-46 (3.2e-72, 2.9e-20) | | VEGFA | 1.1e-33 (2.6e-53, 4.8e-14) | |

**AIC: Akaike information criterion**

***Estimates are in ng/ml scale.**

****Peak severity on an ordinal scale with outpatient-level= 0, inpatient-level=1, ICU-level care =2, death =3**

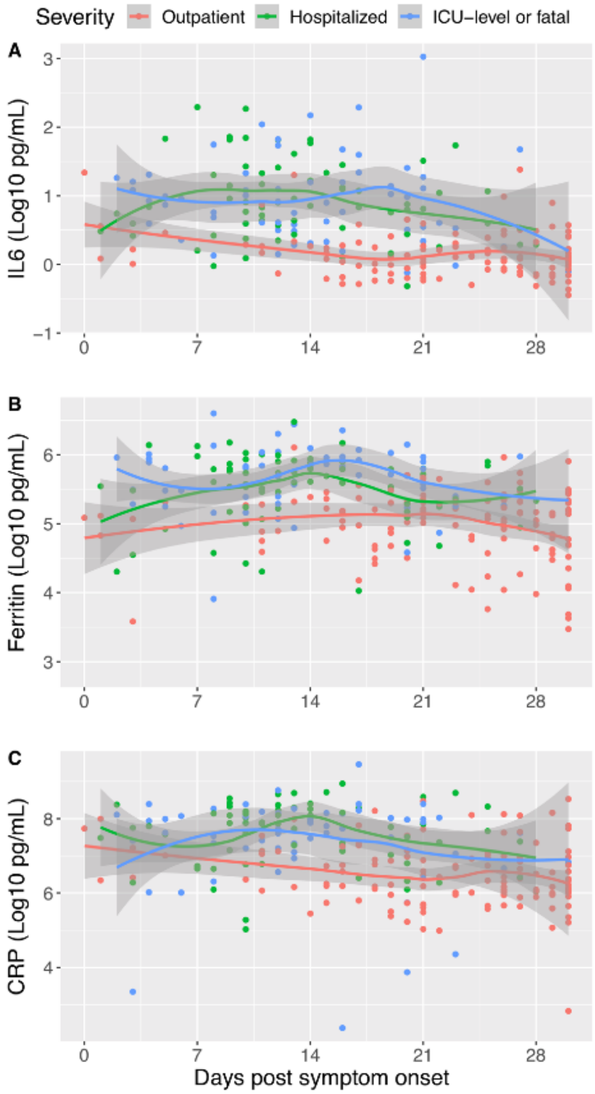


**Supplementary Figure S1.** Scatter plot with plasma IL6 (A), ferritin (B), and CRP (C) over time with LOESS (locally estimated scatterplot smoothing) curves stratified by peak severity from EPICC cohort between 0 to 29 days post symptom onset. While participants with mild illness have a down sloping LOESS curve, participants with moderate or severe illness had a late peaking curve during the third and fourth week of illness (the inflammatory phase). Each point represents a sample level with some participants having multiple samples collected. Points are jittered to avoid overplotting.

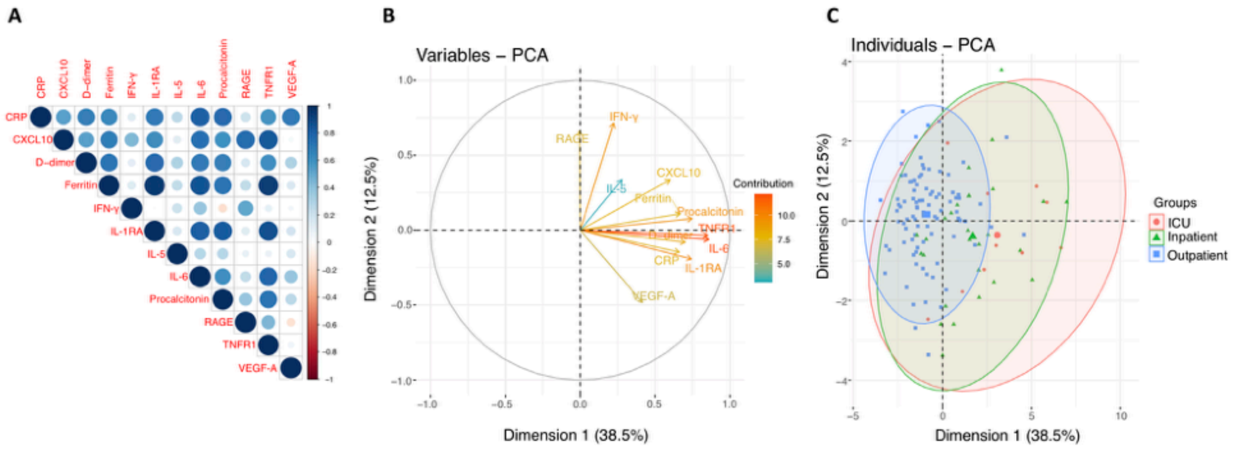


**Supplementary Figure S2.**A: Spearman's correlation matrix of inflammation biomarkers with size and color scale for correlation coefficient; B: PCA showing dimensionality of different analytes; C: PCA showing dimensionality of different analytes with peak severity labeled by color.

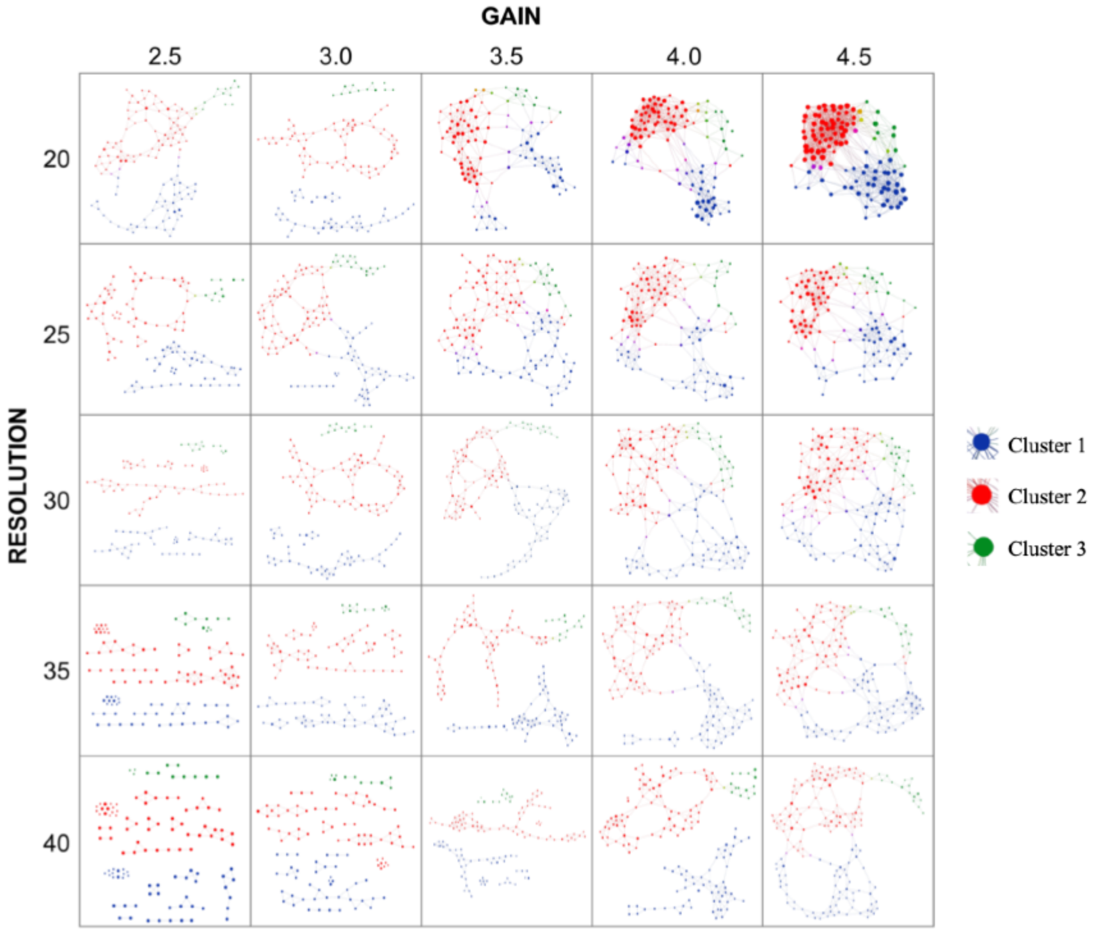


**Supplementary Figure S3.** TDA networks were generated for a range of resolution settings to examine the persistence of subject clusters and their interrelatedness. Clusters 1 (blue), 2 (red), and 3 (green) were observed consistently throughout different resolution and gain settings in the EurekaAI Workbench platform (SymphonyAI, Los Altos, CA, USA). Each node represents a combination of 12 plasma protein analyte levels and its size increases with the number of participants that are included.

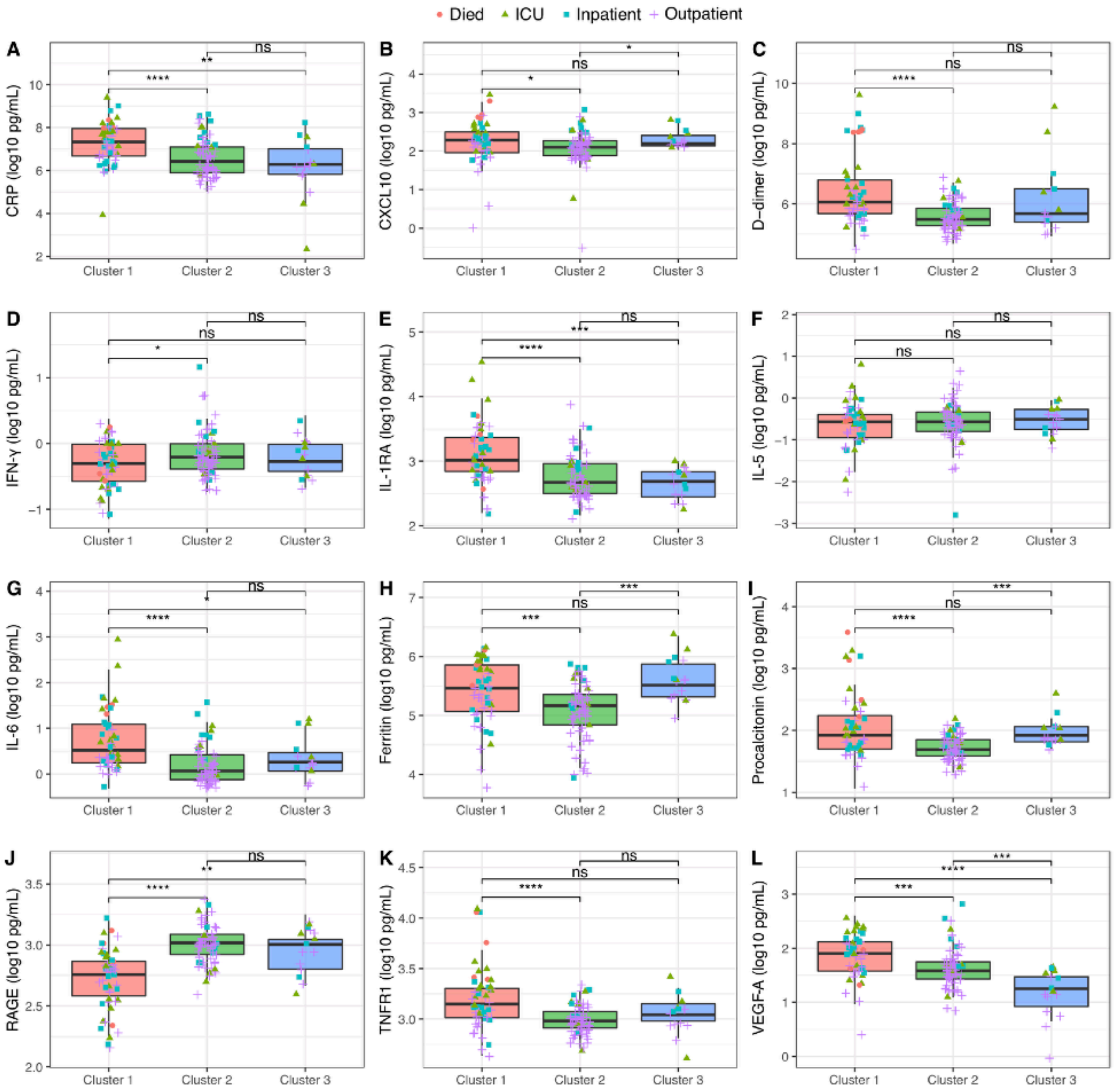


**Supplementary Figure S4.** Immune biomarkers divided by TDA cluster. Each sample with peak COVID severity labeled by color and shape. Kruskal-Wallis test performed comparing analyte levels between clusters. *: $p \leq 0.05$; **: $p \leq 0.01$; ***: $p \leq 0.001$; ****: $p \leq 0.0001$
